# A Novel *de novo TP63* Mutation in Whole Exome Sequencing of a Syrian Family with Oral Cleft and Ectrodactyly

**DOI:** 10.1101/2022.02.10.21265142

**Authors:** Claire L. Simpson, Danielle C. Kimble, Settara C. Chandrasekharappa, NISC Comparative Sequencing Program, Khalid Alqosayer, Emily Holzinger, Blake Carrington, John McElderry, Raman Sood, Ghaith Al-Souqi, Hasan Albacha-Hejazi, Joan E. Bailey-Wilson

## Abstract

Oral clefts and ectrodactyly are common, heterogeneous birth defects. We performed whole exome sequencing (WES) analysis in a Syrian family. The proband presented with both orofacial clefting and ectrodactyly. A paternal side second-degree relative with only an oral cleft was deceased and unavailable for analysis. Variant annotation, Mendelian inconsistencies, and novel variants in known cleft genes were examined. Candidate variants were validated using Sanger sequencing. Twentyeight candidate *de novo* events were identified, one of which is in a known oral cleft and ectrodactyly gene, *TP63* (c.956G>T, p.Arg319Leu), and confirmed by Sanger sequencing. A triallelic SNV was also found in *HLA-DRB5*, which is similar to and overlaps the known oral cleft gene *HLA-DRB1*.

*TP63* mutations are associated with multiple autosomal dominant orofacial clefting and limb malformation disorders. The p.Arg319Leu mutation seen in this patient is *de novo* but also novel. A known mutation 1bp upstream (rs121908839, c.955C>T, p.Arg319Cys) causes ectrodactyly, providing evidence that mutating this codon is deleterious. It is unclear whether this patient’s mutation is responsible for the entire phenotype. Generation and characterization of *tp63* knockout zebrafish showed necrosis and rupture of the head at 3 days postfertilization. The embryonic phenotype could not be rescued by injection of zebrafish or human mRNA. Further functional analysis is needed to determine what proportion of the phenotype is due to this mutation.

## Introduction

In mammalian development, the head is one of the most complex structures to form. The tissues develop from endoderm, mesoderm, ectoderm, and cranial neural crest cells, and the regulation of growth and differentiation is controlled by signaling between different cellular components both spatially and temporally in a highly complex process that can be easily disrupted. This intricate interplay of numerous factors means that cleft lip, with or without cleft palate, is a clinically and genetically heterogeneous trait with multiple genes and regions mapped. There are over 400 syndromes that have oro-facial clefts as a key feature (1), but the majority of patients born with an oral cleft are non-syndromic, where the cleft is the only malformation in the child. Only a small proportion of causal genes have been identified for either syndromic or non-syndromic oral clefts.

Ectrodactyly (also known as split hand/split foot malformation, SHFM) is another clinically heterogeneous malformation(2, 3) in which the absence of the central rays produces a deep median cleft in the autopod, one of the skeletal elements of the developing limb. Like oral clefts, ectrodactyly can occur as an isolated entity or as part of a syndrome. During development, gradients of signaling molecules in three spatial directions control the patterning of the limbs. Three specialized cell clusters control this process through differentiation and proliferation; the apical ectodermal ridge (AER), the progress zone, and the zone of polarizing activity. Both genetic and environmental risk factors are known to disrupt the function of the AER and cause ectrodactyly. Mutations in *TP63*(4–7) and WNT10B(8) have been associated with ectrodactyly, and other regions of the genome have been mapped as containing some still unidentified causal genes. Duplication of 10q24 is also associated with ectrodactyly and is the most common cause of SHFM in humans, accounting for approximately 20% of cases (9).

Whole exome sequencing (WES) has successfully identified the causal variants in a range of Mendelian diseases. Here we present the results of a WES study of a single Syrian family with a child affected with both orofacial cleft and ectrodactyly.

## Materials and Methods

### Recruitment and Clinical Features

A collaborative study of familial orofacial clefts, with an emphasis on nonsyndromic oral clefts, was instituted in the Syrian Arab Republic in 1998 by investigators at the National Human Genome Research Institute, National Institutes of Health, USA, and clinicians at the Ibn Al-Nafees Hospital, Damascus, Syria(10, 11). Families were ascertained through at least one individual affected with non-syndromic cleft lip with or without cleft palate. Of these families, those with two or more family members affected with orofacial clefts were invited to enroll in this genetic study. The study was approved by the Institutional Review/Ethics Boards of the National Human Genome Research Institute, NIH (USA), and the Ibn Al-Nafees Hospital (Damascus, Syrian Arab Republic). All study participants provided written informed consent (in Arabic), and the study followed the tenets of the Declaration of Helsinki.

The informed consent forms and the protocol on file with the Institutional Review Board at the NIH both guarantee the pedigrees will never be published to protect the privacy of the study participants because these pedigrees are readily identifiable given the rarity of such multiplex oral cleft families. Therefore, only a redacted and disguised version of the pedigree is shown here. Subjects enrolled in this study were all examined by the same local physicians and were subjected to standardized interviews. The ascertainment of all families followed the clinical guidelines proposed by the International Consortium for Oral Clefts Genetics(12). During the enrollment of these families, one family was identified with one individual who was affected with non-syndromic bilateral cleft lip and palate (deceased) who had a relative with bilateral cleft lip and palate as well as ectrodactyly (bilateral split hand and split foot malformation). The current study involves this specific family. The paternal second-degree relative with cleft lip and palate, who died in childhood, was reported to have no other clinical abnormalities; however, he was not available for examination. Upon clinical examination, the affected relative was found not to have any symptoms of ectodermal dysplasia, but he did have redness of his eyes and reported frequent, chronic tearing. He was developmentally normal and had no other apparent clinical symptoms. The deceased affected relative had between 10 and 15 unaffected siblings (including the affected relative ‘s father, the exact number of siblings disguised to protect the family ‘s privacy). The affected relative had between 6 and 10 unaffected siblings (exact number disguised to protect the family ‘s privacy). The affected proband ‘s parents were reported to be related, but the family did not specify the exact relationship of the proband ‘s parents.

### Whole Exome Sequencing (WES)

Genomic DNA was extracted from EDTA-treated blood, as described by Bellus *et al*.(13). WES was performed in the patient plus his parents and two unaffected siblings using the TruSeq DNA Sample Preparation v2 method (Illumina, San Diego), followed with Illumina ‘s TruSeq Exome Enrichment Kit protocol and sequenced using the Illumina HiSeq2000 with version 3 chemistry to a depth of at least 40 million paired-end 100 base reads for each sample. Image analysis and base calling were performed with default parameters using Illumina Genome Analyzer Pipeline software (RTA version 1.17.20 and CASAVA 1.8.2).

### Alignment and Genotype Calling

Reads were mapped to NCBI build 37 (hg19) with Novoalign V2.08.02. The aligned lane bam files were merged, sorted, and indexed. Duplicate sequence reads derived from the same original DNA molecule, a PCR artifact characterized by molecules having the exact same alignment coordinates for both Read 1 and Read 2, were removed with Samtools. These alignments were stored in BAM format and then fed as input to bam2mpg (http://research.nhgri.nih.gov/software/bam2mpg/index.shtml), which called genotypes at all covered positions using a probabilistic Bayesian algorithm (Most Probable Genotype, or MPG). These genotype calls have been compared against Illumina Human 1M-Quad genotype chips, and genotypes with an MPG score of 10 or greater showed >99.89% concordance with SNP Chip data. Sequence bases with a quality score less than 20 (Q20) were ignored. Only reads with mapping quality greater than 30 were included for the variant calling.

### Post-Calling Quality Control

Genotypes were zeroed out for read depth < 10, genotype quality [GQ] < 10 or a GQ to read depth ratio of < 0.5 in Golden Helix SVS v7. Mendelian inconsistencies were identified and examined as candidate *de novo* mutations. Candidate recessive genes were identified by classifying all variants which were heterozygous in both parents and homozygous in the patient but heterozygous or homozygous for the common allele in the other unaffected offspring.

### Annotation

The variants were annotated using Annovar (http://annovar.openbioinformatics.org/en/latest/user-guide/gene/). Several filtering and prioritization steps were applied to reduce the number and identify potentially pathogenic mutations, similar to the methods used in previous studies(14, 15). Missense variants were sorted by the degree of severity of functional disruption prediction using CDPred. Variants detected in dbSNP (version 137), 1000 Genomes, NHLBI 6500ESP and HGMD were annotated. Detection of Candidate Recessive Loci Candidate recessive loci were identified using custom scripts in R (https://www.r-project.org/) and available from the authors on request. Briefly, the script filters loci based on mean allele frequencies from 1000 Genomes, NHLBI ‘s Exome Sequencing Project (ESP), and the Exome Aggregation Consortium (ExAC) combined, identify all loci heterozygous in both parents and homozygous for the rare allele in the patient and then filtered this list to remove variants homozygous for the rare allele in unaffected siblings of the patient.

### Detection of Mendelian Inconsistencies

Mendelian error detection was performed in PLINK(16), and candidate variants were examined in GoldenHelix SVS.

### Sanger Sequencing

To confirm candidate variants of interest detected by the analyses above, Sanger sequencing was performed on two variants in 7 individuals, consisting of the patient (affected with bilateral cleft lip and palate and ectrodactyly), his parents, and four unaffected siblings. Primers were designed with M13 tags attached for the regions of interest in *TP63* and *HLA-DRB5* (Supplementary Table S1). PCR products were generated using the KAPA2G Fast HotStart ReadyMix kit (KAPA Biosystems), 2µM primer, and 2.5ng genomic DNA. PCR products were treated with ExoSAP-IT (Affymetrix), and these treated products were used in BigDye Terminator v3.1 Cycle Sequencing reactions (Applied Biosystems) with 10µM M13 forward and reverse primers, followed by Sanger sequencing on an ABI 3730xl DNA Analyzer (Applied Biosystems). Sequence tracings were analyzed with Sequencher (Gene Codes) software.

### Allele-Specific Cloning

Due to the difficulty in determining the correct genotype in the patient in the *HLA-DRB5* sequence data, allele-specific cloning and sequencing were performed in the patient and both parents. The CloneJET PCR Cloning kit (ThermoScientific) was used to ligate PCR products into the pJET1.2 vector, and they were transformed using competent E. coli cells. Direct PCR of colonies was performed with the same *HLA-DRB5* primer set used in the original PCR, followed by ExoSAP-IT treatment, BigDye reactions, and Sanger sequencing as detailed in the section above.

### Zebrafish husbandry and Ethics statement

All zebrafish experiments were performed in compliance with the National Institutes of Health guidelines for animal handling and research using an Animal Care and Use Committee (ACUC) approved protocol G-05-5 assigned to RS. Wildtype (WT) zebrafish strain TAB5 was used for all experiments. Zebrafish husbandry and embryo staging were performed as described previously(17).

### Generation of *tp63* mutants and genotyping

Two single guide RNAs (sgRNAs) targeting exons 5 (GGATG-GCAGGTGATGGAGAG) and 6 (GTATGACTGCACC-CTGGGGT) of *tp63* (Ensembl transcript ID: ENS-DART00000127965.4) were designed using the ‘ZebrafishGenomics ‘ track on the UCSC Genome Browser. Synthesis of target oligonucleotides (Integrated DNA Technologies), preparation of mRNA, microinjections, CRISPR-STAT to evaluate sgRNA activity, and mutant generation were carried out as described previously(18–20). Primers used for screening and genotyping by fluorescent PCR were as follows: E5-Fwd (5 ‘-GCTTCTCAACAGCATG-GATC) and E5-Rev (5 ‘-TCCAGGTTGCAGATTTGGC), E6-Fwd (5 ‘-CTCCACAGAGTTGAAGAAGC), and E6-Rev (5 ‘-CATTGAACTCTCTGCTCAGC). M13F adapter (5 ‘-TGTAAAACGACGGCCAGT) was added to the 5 ‘ end of each forward primer, and PIG-tail (5 ‘-GTGTCTT) was added to the 5 ‘ end of each reverse primer as described(21).

### Time lapse imaging

Embryos were immobilized in 1x tricaine and images were acquired every 5 minutes for a 24 hour period using a Leica M125 microscope equipped with an MC170HD camera and the Leica Application Suite (LAS) V4.4. Post-processing of images was done within the LAS software.

### Microinjections of mRNA for rescue of phenotype

The following clones were obtained in pBluescriptII for zebrafish *tp63* (NM_152986.1) and human *TP63* (NM_003722.4) (Genescript). Plasmid DNA was digested with XhoI and mRNA was synthesized using a T7 mMessage mMachine kit (Ambion). Following transcription, polyA tailing was performed, and RNA was purified by LiCl precipitation. Injection of mRNA (100pg to 1ng) into WT embryos was done to determine the appropriate dose. Injections (500pg) of human or zebrafish mRNA were then performed in embryos from in-crosses of *tp63*^*+/-*^ fish. Embryos were observed at 72 hours post-fertilization (hpf) and 78 hpf for phenotype and collected for genotyping.

## Results

### Data Quality

Before quality control, there were a total of 185,474 SNVs and 18,058 INDELs. After applying quality control filters, variants were dropped for being monomorphic or where all individuals were heterozygous. There were 11,905 INDELS and 137,989 SNVs available for analysis after all filtering steps. Using a mean allele frequency calculated from the 1000 Genomes Project, the NHLBI Exome Sequencing Project, and the Exome Aggregation Consortium, loci were filtered and were retained only if their mean minor allele frequency was less than or equal to 10%.

### Selection of Candidate Genes and Validation by Sanger Sequencing

Thirty-four loci were homozygous for the minor allele in the patient but heterozygous in the parents and homozygous for the major allele or heterozygous in the unaffected siblings. None of these 34 variants were in known cleft genes, and none of the genes identified were good candidates for orofacial clefting by functionality. The complete list of variants can be found in Supplementary Table S2. Examining Mendelian inconsistencies revealed 28 candidate *de novo* events, of which only two were well supported by examination of the alignment and had enough biological plausibility for follow-up. Details of all 28 loci are listed in Table 1.

**Table 1.**
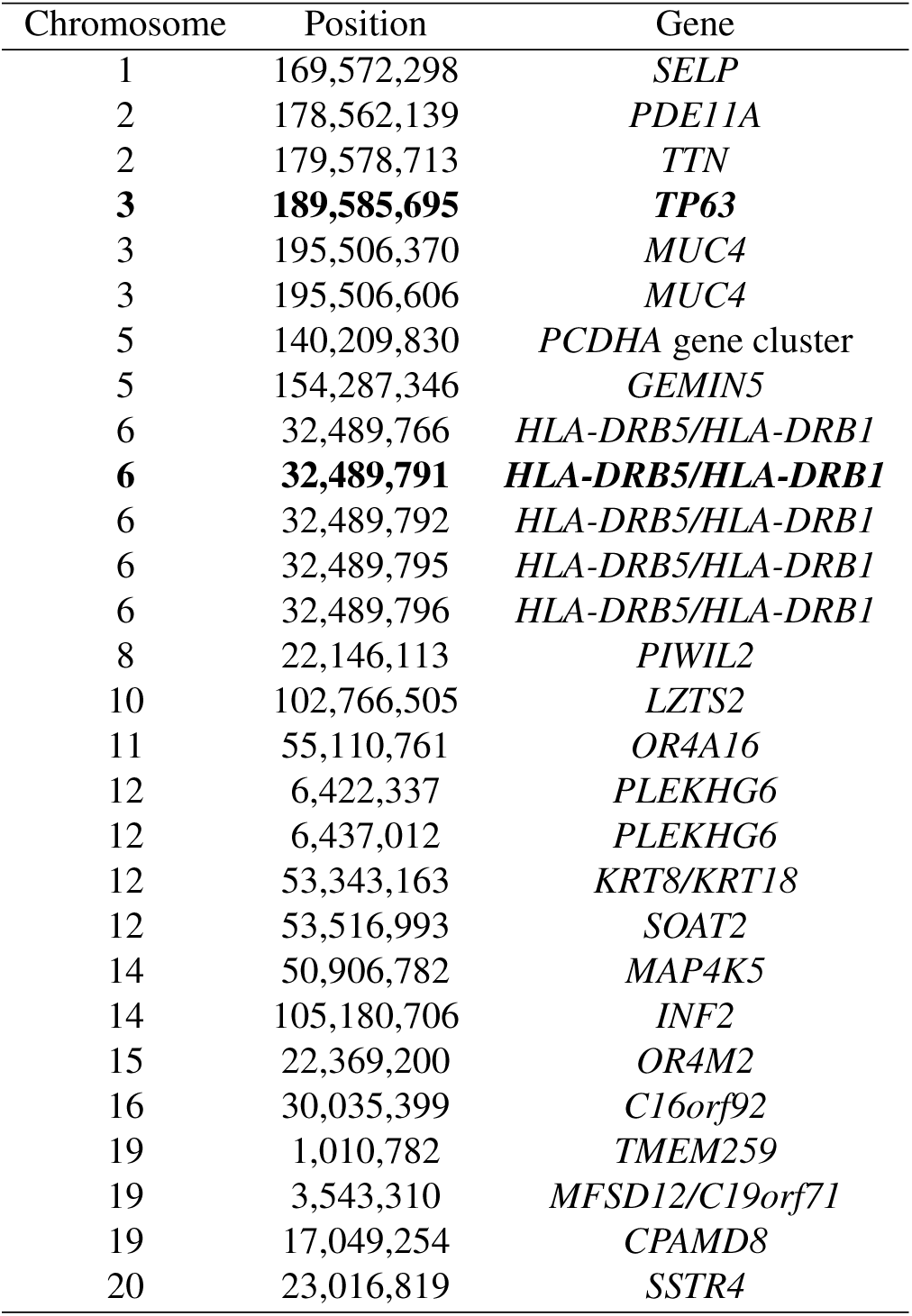
Candidate *de novo* mutations identified in the patient’s whole exome sequence data, with the two variants selected for follow-up in bold text.

The most interesting of the *de novo* candidates was a nonsynonymous, single base substitution in *TP63* (c.956G>T, p.Arg319Leu). Examination of the reads in Golden Helix Genome Browse showed that the call had good coverage and was present in reads in both directions, consistent with a heterozygous mutation. Sanger sequencing in the entire pedigree confirmed this was a likely true *de novo* mutation (Figure 1). Examination of whole genome or whole exome sequencing of 37 additional individuals with non-syndromic oral clefts from other Syrian oral cleft families(22) revealed only two coding variants in *TP63*; rs140508531, a rare synonymous SNV previously seen in ExAC and ESP. The other was a synonymous SNV not reported in any databases. Genotypes for this variant for each individual called from the Sanger sequencing can be found in Table 2. We also examined exome and genome sequences from other populations of non-syndromic oral cleft patients (populations details and numbers can be found in Table 3 and found two more synonymous variants in *TP63*; one had never been seen in any of the online databases, the other was rs33979049, an uncommon SNV seen in 1000 Genomes at a MAF of between 1 and 5%, depending on population.

**Fig. 1.**
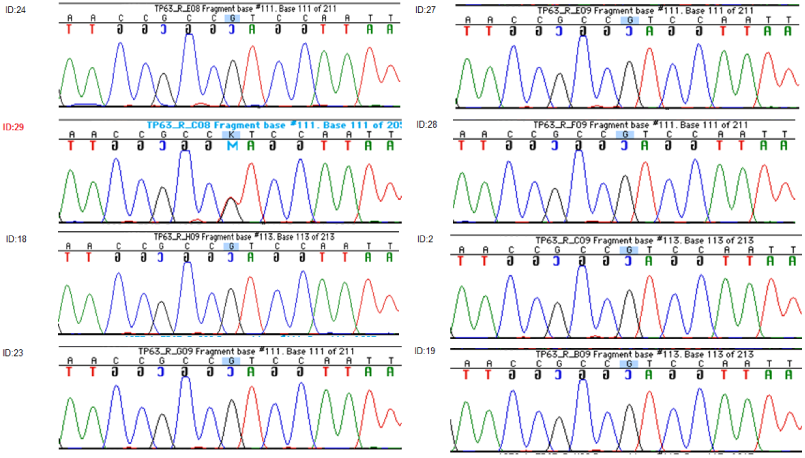
Sanger sequencing results of *TP63* mutation in all available family members.

**Table 2.**
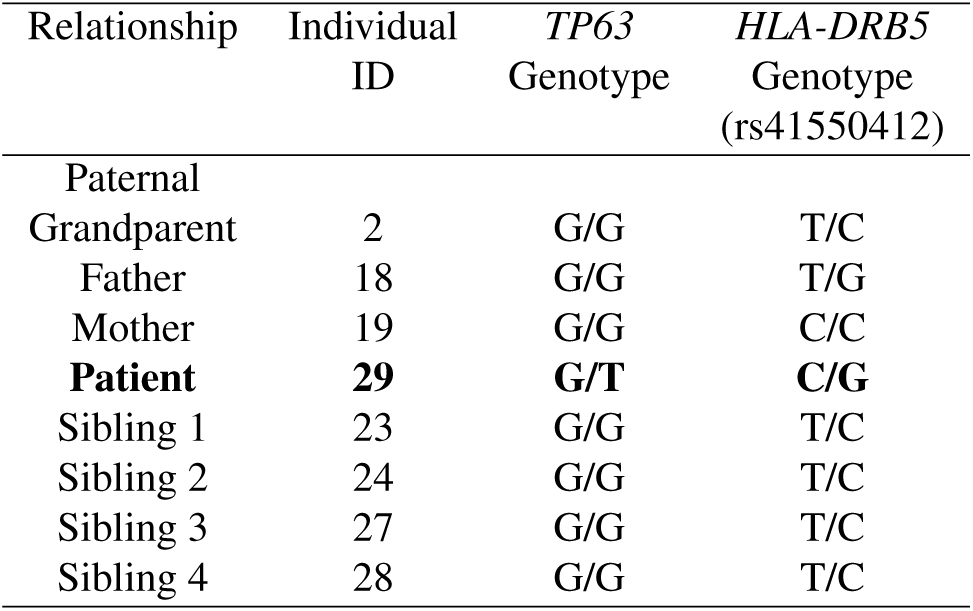
Genotype results of *TP63* and *HLA-DRB5* Sanger sequencing, with the patient highlighted in bold text.

**Table 3.** Genotype results of *TP63* and *HLA-DRB5* Sanger sequencing, with the patient highlighted in bold text.

| Population | Individuals (families) |  |
| --- | --- | --- |
|  | with WES** | with WGS** |
| Syrian | 16 (8) | 37 (14) |
| Indian | 26 (12) | 37 (19) |
| Filipino | 22 (11) | 78 (18) |
| German | 38 (19) | 0 |
| Taiwanese | 2 (1) | 3 (1) |
| European-American | 3 (1) | 4 (1) |
| Chinese | 2(1) |  |
| Guatemalan | 3 (2) |  |
| Singaporean | 5 (3) |  |

The second putative *de novo* candidate was a third allele in the known SNP, rs41550412 in *HLA-DRB5*. Annotation of the variant in dbSNP and examination of the reads in Genome Browse suggested that this was a tri-allelic SNP and might be segregating normally in the family. Sanger sequencing confirmed this hypothesis (Table 2) and showed this family was segregating four additional non-synonymous variants in this gene (Figure 2) that had not been correctly captured by the WES due to the coverage level and allele-specific read imbalance at this location. Allele-specific cloning and Sanger sequencing of both parents and the patient was performed to clarify the inheritance of 5 loci in this gene (Figure 3, Supplementary Table S3). The patient had a unique combination of all five variants (Figure 2 and Figure 3) that any of his 4 genotyped unaffected siblings did not share. However, none of these variants are rare, so it is also possible this is a chance finding. However, compared to the other non-syndromic oral cleft sequence data we have generated (Table 3), none of the additional exome or genome sequences examined shared any of these variants.

**Fig. 2.**
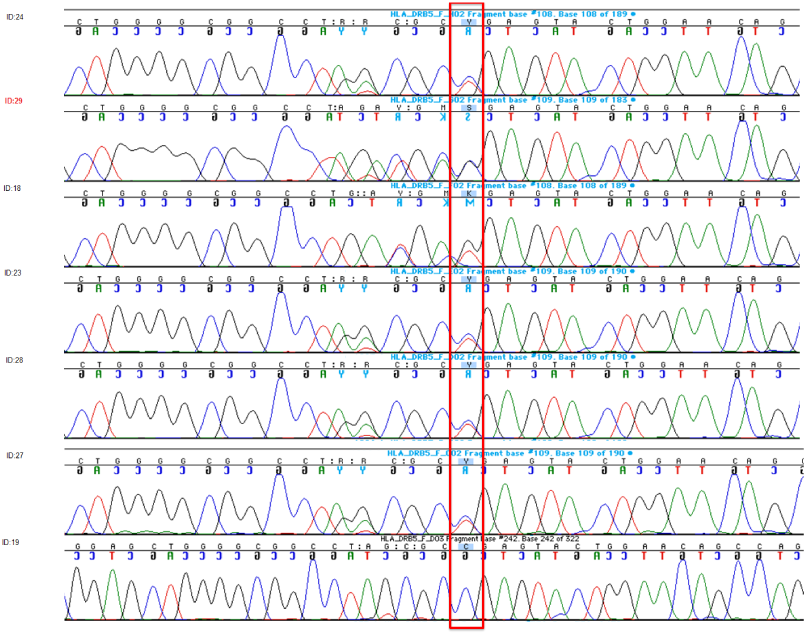
Sanger sequencing results of *HLA-DRB5* variant in all available family members

**Fig. 3.**
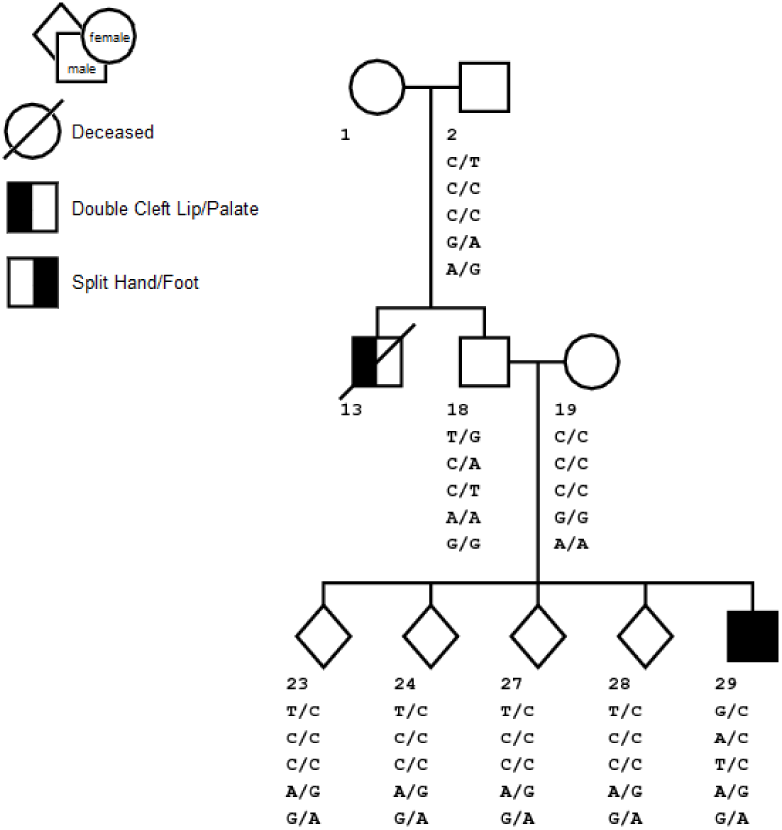
HLA-DRB5 Genotypes from Sanger Sequencing for all available family members

We also examined 67 candidates genes (Supplementary Table S4) for recessively inherited variants in this family. However, no coding variation in these genes produced unique genotypes in the patient compared to his siblings.

### Generation of *tp63*^*-/-*^ Zebrafish Mutants

To validate the role of Tp63 in this proband ‘s phenotype, we generated *tp63* knockout fish lines using CRISPR/Cas9 technology. To ensure that the mutation affects all known isoforms, we selected gRNAs to exons 5 and 6 that are common to all isoforms (Supplemental Figure 1). Two mutant alleles (del 5bp in exon 5 and del 2bp in exon 6) predicted to cause frameshifts with premature truncations of the protein, were selected for the study (Figure 4A). These mutant alleles have been given designations from the Zebrafish international Resource Center (http://zfin.org/action/feature/line-designations) as *tp63*^*hg118*^ (c.358_362delTCTCC; p.Ser120Ilefs*17) and *tp63*^*hg119*^ (c.540_541delCC; p.Gln181Glyfs*11).

**Fig. 4.**
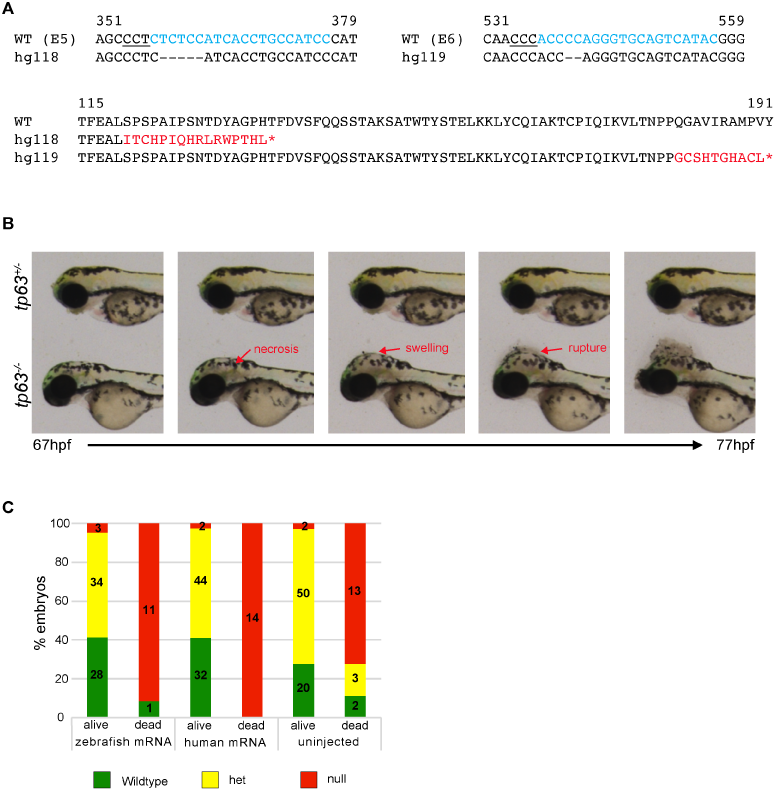
Generation and characterization of *tp63* knockout mutants. (A) Top panel: Nucleotide sequences of CRISPR target regions in exons 5 and 6 with sgRNAs marked in cyan, PAM sites underlined, and deleted nucleotides in the mutant alleles marked by dashes. Bottom panel: An alignment of WT and mutant proteins with the frameshift and premature stop codons marked in red. (B) Phenotype of the *tp63*^*-/-*^ embryos shown as images at different time points from a time lapse video taken from 67 to 77 hpf. Images of a heterozygote sibling are shown for control. At 67 hpf the *tp63*^*-/-*^ embryo is indistinguishable from its heterozygous sibling. Shortly after necrosis within the head begins followed by swelling and rupture as marked by red arrows. (C) Data from rescue experiments by injections of 500 pg of zebrafish or human *tp63* mRNA shown as bar graphs. Embryos were observed for phenotype between 72 – 78 hpf, separated into dead or alive groups and genotyped. Different groups of embryos that were scored are shown on the X-axis and percentages of embryos of different genotypes are shown on the Y-axis.

### Loss of *tp63* Leads to Necrosis and Rupture of Head at 3 dpf

To determine the effect of loss of function of *tp63* on development, we performed pairwise crosses of heterozygous fish for each mutant allele separately and observed their progeny for morphological phenotypes. Homozygous mutant embryos for both alleles displayed similar morphological phenotype and hence are collectively referred to as *tp63*^*-/-*^. The *tp63*^*-/-*^ embryos were indistinguishable from their wildtype (WT) and heterozygous clutch mates for the first two days of development. At 3 days post fertilization (dpf), they displayed a rupture of the head followed by death (Figure 4B). Time lapse imaging revealed that about 2-3 hours prior to head rupture, necrosis and swelling occurs in the head region of mutant embryos (Figure 4B). Therefore, we were unable to evaluate jaw and fin development in *tp63*^*-/-*^embryos to validate its role in the patient ‘s phenotype.

### Rescue of embryonic phenotype by injection of zebrafish or human Mrna

We hypothesized that if the *tp63*^*-/-*^ phenotype can be rescued by WT *tp63* mRNA, then we could evaluate the patient variant for its effect on Tp63 function using the rescue assay. We performed dose response curve by injecting either human or zebrafish WT *tp63* mRNAs into WT embryos and monitoring their viability and morphological phenotype (data not shown). Subsequently, 500pg of zebrafish or human mRNA was injected into embryos from an in-cross of *tp63*^*hg118/hg118*^ fish. We did not observe any phenotypic or survival improvements in the injected *tp63*^*hg118/hg118*^ embryos (Figure 4C), indicating that the phenotype is too severe to be rescued by complementation by WT mRNA. Therefore, the rescue assay was not applicable for validation of the patient variant.

## Discussion

Mutations in *TP63* are known to cause a number of different malformation syndromes, which include orofacial clefting or limb malformations in their phenotypic presentation, including Ectrodactyly, Ectodermal Dysplasia and Cleft Lip/Palate Syndrome 3 (EEC3), Split Hand/Foot Malformation 4 (SHFM4), Hay Wells Syndrome and Acro-dermatoungual-lacrimal-tooth (ADULT) syndrome. Most of the known mutations in this gene are dominant. Although this mutation appears to be novel, with no recorded instances in public variation or clinical databases, the *SHFM4* mutation 1bp upstream (rs121908839, c.955C>T, p.Arg319Cys) affects the same codon. However, it produces a different amino acid change providing strong evidence that mutating this amino acid is deleterious. These mutations in exon 7 of *TP63* are in the DNA binding domain of the protein and are highly evolutionarily conserved. However, it is not clear whether this mutation is only responsible for the ectrodactyly or whether it is also responsible for the oral cleft seen in the patient. The p.Arg319Cys mutation is so far known only to cause ectrodactyly(23) and not orofacial clefting, and so it cannot be ruled out that the patient has compound mutations in different genes causing the oral cleft and ectrodactyly phenotypes. There is considerable heterogeneity in the phenotypic presentation of *TP63* mutations, as recently demonstrated by Alves *et al*.(24), so either possibility cannot be easily ruled out. Although *HLA-DRB5* has not been associated with orofacial clefting or limb malformations, it is similar to the known oral cleft gene *HLA-DRB1*, some transcripts of which may overlap. This made it of particular interest, especially considering it cannot be ruled out that the *TP63* mutation may not be responsible for the oral cleft part of the phenotype, given the affected relative and the consanguinity of the patient ‘s parents. The patient is a unique compound heterozygote at 4 locations in the *HLA-DRB5* gene, and further study of this gene both *in vitro* and *in vivo* may be warranted to determine if this variant contributes to the phenotype in the patient. Whole exome sequencing has been used to great effect to find causal mutations in patients with genetically heterogeneous diseases. Our study has identified a previously unknown mutation in a gene known to be associated with both oral clefts and ectrodactyly. However, sequencing alone cannot determine exactly which this mutation causes parts of the observed phenotype. It is possible that the orofacial cleft may have another etiology since the patient ‘s relative also had an oral cleft independent of the *de novo TP63* mutation seen here. However, given the prevalence of oral clefts in the Syrian population and the known consanguinity in this pedigree, the relative ‘s cleft may be etiologically distinct from his proband ‘s. Unfortunately, because the second degree relative died in childhood, determining causation becomes more complex. The results from our zebrafish experiments were intriguing if not definitive. We were unable to do functional validation of the patient variant due to severity of the knockout phenotype and inability to rescue it by mRNA injections. A recent study has demonstrated that *tp63* is required for ectoderm specification during zebrafish development(21). Interestingly, *tp63* mutant embryos used in this study died prior to the stage at which head necrosis was observed in our mutant lines (25). These phenotypic differences between different mutant alleles are most likely due to their effects on the multiple isoforms of *tp63*. In-depth functional analysis is both expensive and time-consuming, but there is only so far that computational and statistical methods can take us and additional in vivo studies in model animals may be required. This study represents the first study of its kind in Syria. The family structures in this region can be challenging and the incidence of oral clefts is high. Studying oral clefts is of great significance to global health because it is common. Whereas corrective surgery in early childhood can mitigate most of the effects of the disorder, this surgery is out of reach for many living in low- and middle-income countries, to devastating effect. Understanding the ways in which genes influence the development of the trait is an important step to determining mechanisms and potential preventive options in the future for individuals at high risk of having a child with an oral cleft. Even *de novo* variants as presented here can provide those insights.

## Data Availability

Some data produced in the present work are contained in the manuscript. Patients were not consented for broad data sharing and ethical concerns over identification of families means some data cannot be shared.

## ACKNOWLEDGEMENTS

We would like to express our gratitude to the families who participated in this study. The authors would like to acknowledge Drs. Mary Marazita, Jeff Murray, Elisabeth Mangold, Alan Scott, Ingo Ruczinski, and Terri Beatty for contributing lookup of variants in our genes of interest in their sequence data from multiplex non-syndromic oral clefts families. This work was supported in part by the Intramural Program of the National Human Genome Research Institute, National Institutes of Health, and grants R01 DE014581 (International Genetic Epidemiology of Oral Clefts) and U01 DE020073 (Oral Clefts: Moving from Genome-Wide Studies Toward Functional Genomics). This work utilized the computational resources of the NIH HPC Biowulf cluster. (http://hpc.nih.gov).

---

**Supplementary Note 1:**
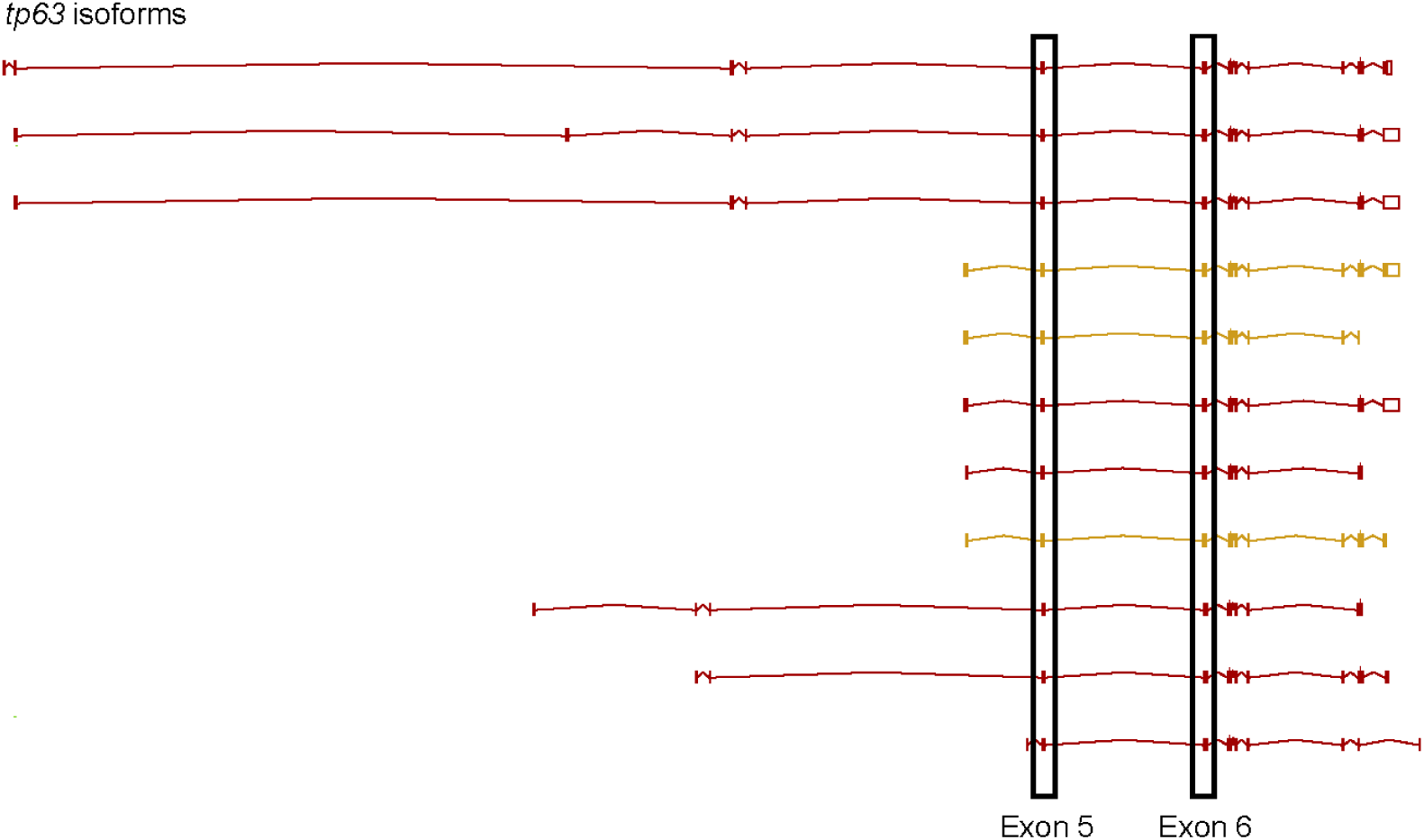
Supplementary Figure 1.

